# An NIH Investment in Health Equity - The Economic Impact of the Flint Center for Health Equity Solutions

**DOI:** 10.1101/2020.10.19.20215137

**Authors:** Cristian I. Meghea, Barrett Wallace Montgomery, Roni Ellington, Ling Wang, Clara Barajas, E. Yvonne Lewis, Sheridan T. Yeary, Laurie A. Van Egeren, Debra Furr-Holden

**Author notes:** **Corresponding author:** Cristian I. Meghea, Michigan State University, Department of Obstetrics, Gynecology and Reproductive Biology, 965 Wilson Road, Room A626B, East Lansing, MI 48824-1316.

## Abstract

**Background:** Health disparities are pervasive and are linked to economic losses in the United States of up to $135 billion per year. The Flint Center for Health Equity Solutions (FCHES) is a Transdisciplinary Collaborative Center for health disparities research funded by the National Institute of Minority Health and Health Disparities (NIMHD). The purpose of this study was to estimate the economic impact of the 5-year NIH investment in FCHES in Genesee County, Michigan.

**Methods:** The estimated impacts of FCHES were calculated using a U.S.-specific input/output (I/O) model, IMPLAN, from IMPLAN Group, LLC., which provides a software system to access geographic specific data regarding economic sector interactions from a variety of sources. This allowed us to model the cross-sector economic activity that occurred throughout Genesee County, Michigan, as a result of the FCHES investment. The overall economic impacts were estimated as the sum of three impact types: 1. Direct (the specific expenditures impact of FCHES and the Scientific Research and Development Services sector); 2. Indirect (the impact on suppliers to FCHES and the Scientific Research and Development Services sector); and 3. Induced (the additional economic impact of the spending of these suppliers and employees in the county economy).

**Results:** The total FCHES investment amounted to approximately $11 million between 2016-2020. Overall, combined direct, indirect, and induced impacts of the total FCHES federal investment in Genesee County included over 161 job-years, over $7.6 million in personal income, and more than $19.2 million in economic output. In addition, this combined economic activity generated close to $2.3 million in state/local and federal tax revenue. The impact multipliers show the ripple effect of the FCHES investment. For example, the overall output of over $19.2 million led to an impact multiplier of 1.75 – every $1 of federal FCHES investment led to an additional $.75 of economic output in Genesee County.

**Conclusions:** The FCHES research funding yields significant U.S. direct economic impacts above and beyond the direct NIH investment of $11 million. The economic impact estimation method may be relevant and generalizable to other TCCs or other large research centers such as FCHES.

## BACKGROUND

Health disparities are pervasive and are linked to economic losses of up to $135 billion per year, including $93 billion in excess medical care costs and $42 billion in missed productivity (Turner, 2018). A Health Inequities Think Tank convened by the National Institutes of Health (NIH) agreed that “confronting health inequities will require engaging multiple disciplines and sectors (including communities), using systems science, and intervening through combinations of individual, family, provider, health system, and community-targeted approaches” (Sampson et al, 2016). The Think Tank issued a set of recommendations for reducing health inequities in the United States, which included embracing broad and inclusive research themes that incorporate multilevel factors; developing research platforms for innovative transdisciplinary research that promote systems science approaches; developing networks of collaborators and stakeholders, and launching transformative studies that can serve as benchmarks; optimizing the use of new data sources, platforms, and natural experiments; and developing unique transdisciplinary training programs to build research capacity.

The National institutes for Health (NIH), through the National institute on Minority health and Health Disparities, established the Transdisciplinary Collaborative Centers for Health Disparities Research Program (TCC). TCC supports regional coalitions of academic institutions, community organizations, health providers and systems, and other stakeholders. Current TCC program grants focus on multilevel chronic disease prevention research, health policy research, social determinants of health, men’s health research, and precision medicine research. As health and disparities are affected by diverse public and private stakeholders, addressing health disparities requires a transdisciplinary approach and strong collaborations between researchers and community organizations, health providers and systems, government agencies, and other stakeholders. Such an approach ensures that findings translate into sustainable changes at multiple levels to reduce inequities in health and improve population health.

The Flint Center for Health Equity Solutions (FCHES) is a Transdisciplinary Collaborative Center (TCC) for health disparities research funded by NIMHD. FCHES focuses its research efforts on health disparities and chronic disease that cross boundaries and directly affect the Flint and Genesee County Community. FCHES is an assembly of stakeholders, including public health researchers, policymakers, health officials, community organizations, and faith-based partners across a range of specialties to mount evidenced-based and promising approaches to prevent chronic disease and reduce health inequities.

Figure 1 presents the structure of FCHES, which includes four cores, two multilevel intervention research projects, and the Flint Area Study, a comprehensive resource of public health data in Flint developed by FCHES. The purpose of this current study was to estimate the economic impact of the 5-year NIH investment in FCHES in Genesee County, Michigan and to provide an economic evaluation model potentially replicable by other TCCs and other large research centers. This study will provide analyses and report on the short-term direct and indirect economic impacts in Genesee County, MI resulting from the expenditures in FCHES; examples and case studies of functional impacts of FCHES; and a brief discussion of the potential medium- and long-term economic impacts of FCHES in other MI counties and statewide. The economic impact is not the only measure of success for FCHES. Another study (Ellington et al, 2020) is presenting the framework for the overall FCHES evaluation.

**Figure 1.**
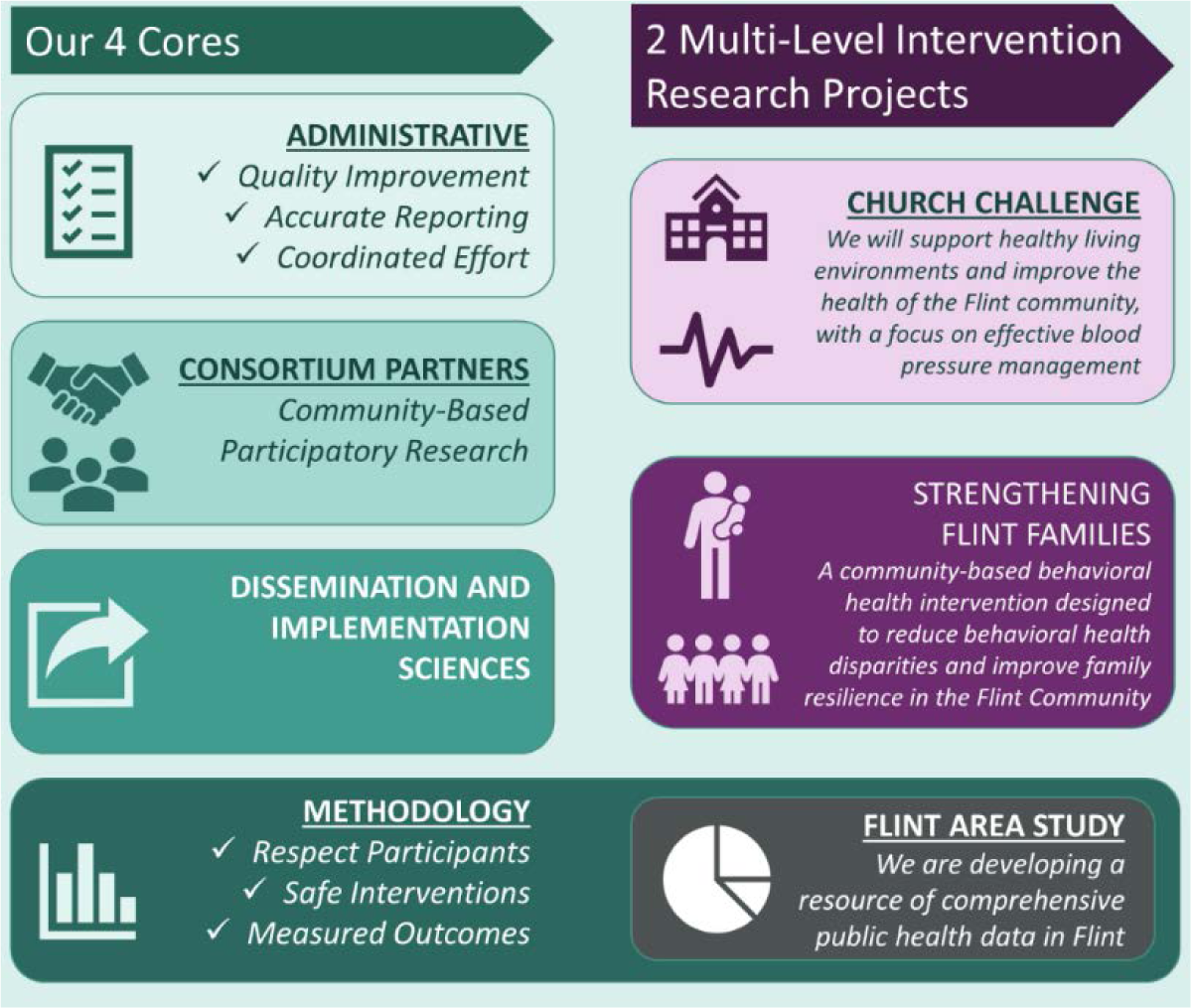
The structure of FCHES – Flint Center for Health Equity Solutions

## METHODS

Figure 2 illustrates the connections between the direct investments in the FCHES and the impacts associated with these investments.

**Fig. 2.**
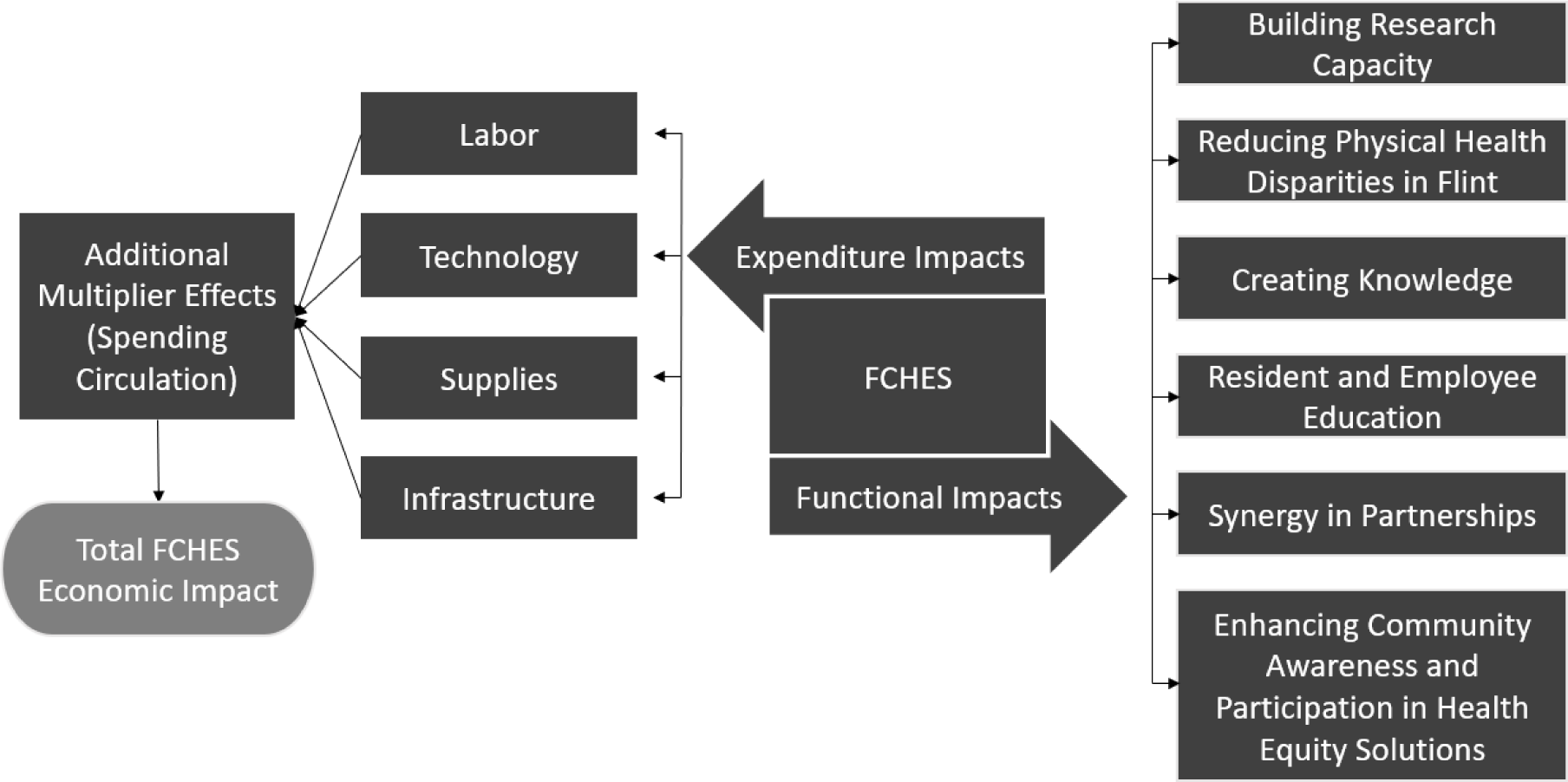
The structure of the impacts associated with FCHES

The economic impact analysis reported in this study uses an input/output (I/O) model to represent the interrelationships among economic sectors (Tuck & Hawkins, 2019; Shrestha & Memhood, 2018). I/O models are quantitative economic models representing the interdependencies between different sectors of a national or regional economy (in this case Genesee County, MI). The estimated impacts of FCHES were calculated using a U.S.-specific I/O model, IMPLAN, from IMPLAN Group, LLC., which provides a software system to access geographic specific data regarding economic sector interactions from a variety of sources. Data from U.S. government sources (e.g. Bureau of Economic Analysis, Department of Agriculture, Bureau of Labor Statistics, Census, and others) are collected and merged on an annual basis to produce a complete set of Social Accounting Matrices (SAMs) for each sector and region of interest. If the data source is not updated annually, the IMPLAN data team estimates the missing pieces and includes this estimate in the annual updates. The SAMs provide a complete picture of an economy and can be used to create a predictive I/O model for estimating economic impacts. In this case, the direct investment from the NIH to the FCHES is plugged into the model as the input and the model specifications and assumptions are used to estimate how that money is spent in Genesee county. This spending on goods and services has a ripple effect in the economy (i.e. multipliers) and the model quantifies the degree to which other sectors and persons in Genesee county benefit from the initial investment. Using data on the spending patterns of 536 individual industry sectors (e.g. different types of farming, construction, manufacturing, and services), we were able to model the cross-sector economic activity that occurred throughout Genesee County as a result of the FCHES investment.

Several considerations went into the IMPLAN model specification. First, due to the nature of the NIMHD investment in research, we selected a single industry sector - Scientific Research and Development Services (IMPLAN sector number 456) – to estimate these economic impacts. Second, the investments are entered into the model in the dollar year of the investment (between 2016 and 2020) and the output results are adjusted to 2020 dollars. IMPLAN computes these based on built in economic “inflators” and “deflators” to allow for the year by year inflation of the U.S. Dollar and inflation or deflation of products, commodities, and services. These year-by-year adjustments allow for the development of a cumulative multi-year impact estimation for the 5 years included in the analysis. Third, our model uses the 2016 year of economic data for the NIMHD investment in 2016, and the 2017 data year for each subsequent annual investment (2017-2020). This is a necessary limitation of the model because the economic data for years 2018 and beyond were not available at the time of analysis. Fourth, because of the nature of scientific research centers and their difference from a typical company is which the owner receives profits, proprietor income is set to $0. Last, we used the actual employment, full time employee percentages (FTE%), and actual compensation of FCHES employees to increase the precision modeling the economic impact of FCHES.

What is commonly referred to as ‘economic impacts’ are calculated in IMPLAN as ‘Output’. The Output metric is composed of 5 categories that represent the ways in which value is added into the economy: 1. Intermediate Expenditures; 2. Employee Compensation; 3. Proprietor Income; 4. Taxes on Production and Imports; 5. Other Property Income. All of these are used to estimate the economic impact within the model. As the IMPLAN model incorporates 536 individual industry sectors, it allows for the modelling of cross-sector economic activity resulting from the FCHES work. The overall economic impacts consist of three impact types: 1. Direct (the specific expenditures impact of FCHES and the sector in question); 2. Indirect (the impact on suppliers to FCHES and the Scientific Research and Development Services sector); and 3. Induced (the additional economic impact of the spending of these suppliers and employees in the economy).

The following results are provided for each impact estimation (see Figure 3): employment, measured in job-years; personal income, including both wages and benefits; value added (analogous to GDP, includes labor income as well as taxes and property income); economic output; state and local tax revenue, including income and property taxes; and federal tax revenue, including contributions to Social Security. An impact multiplier is also provided for each type of data. Multipliers are calculated by dividing the total impact by the direct impact.

**Figure 3.**
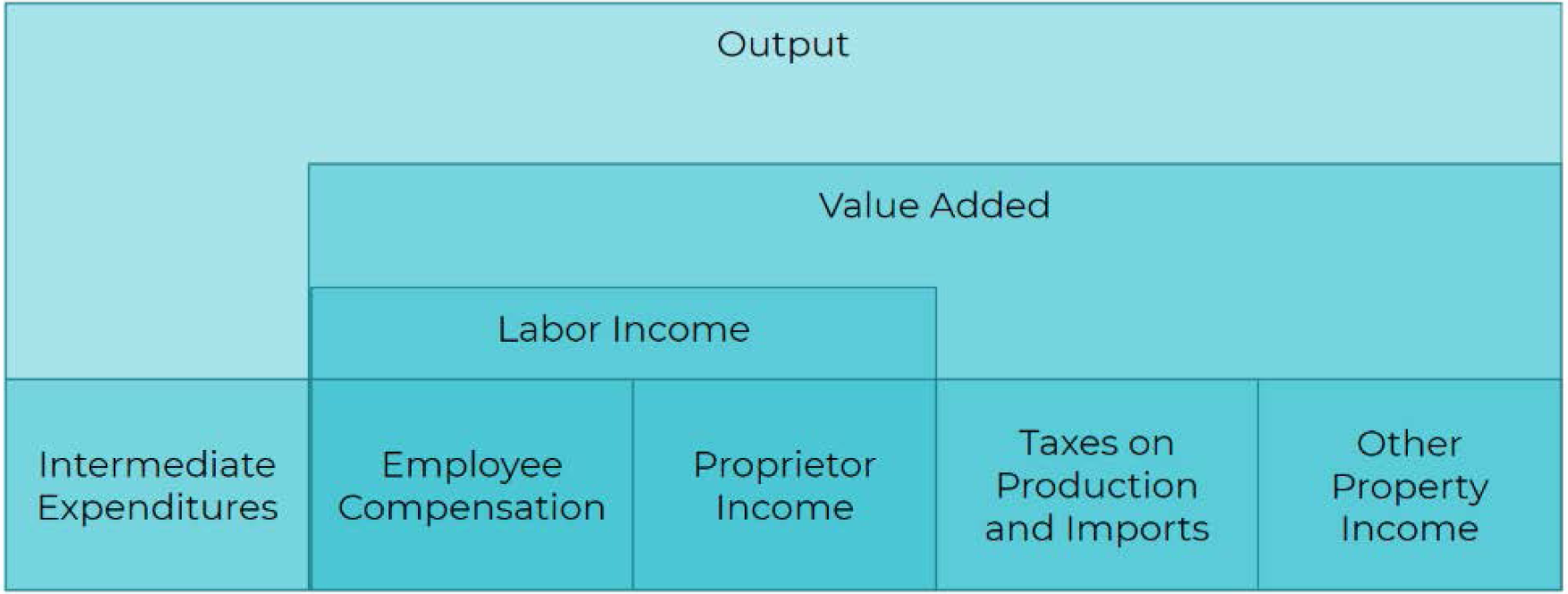
The structure of the IMPLAN economic model (graphic provided by IMPLAN Group, LLC, 2017)

The FCHES research funding data includes the NIH federal investment in FCHES (Table 1). Table 2 reports the impact of one year (2019) of the FCHES funding to enable comparisons to other projects on a year-by-year basis. Table 3 reports the cumulative economic impact of the FCHES federal funding. All amounts are expressed in 2020 equivalent dollars.

**Table 1.** NIH annual investment in FCHES

|  | **2016** | **2017** | **2018** | **2019** | **2020** | **Total** |
| --- | --- | --- | --- | --- | --- | --- |
| *Initial Investment* | \$2,382,051 | \$2,195,822 | \$2,108,424 | \$2,124,467 | \$2,043,062 | \$10,853,827 |
| *Supplemental Investment* | \$0 | \$53,499 | \$110,049 | \$0 | \$0 | \$163,549 |
| **Total** | \$2,382,051 | \$2,249,322 | \$2,218,474 | \$2,124,467 | \$2,043,062 | \$11,017,376 |

**Table 2.** Annual Economic Impact of FCHES Federal Funding in 2019

| ***Impact*** | ***Employment***<br>***(Job-Years)*** | ***Personal***<br>***Income*** | ***Output*** | ***State and***<br>***Local Tax***<br>***Revenue*** | ***Federal Tax***<br>***Revenue*** |
| --- | --- | --- | --- | --- | --- |
| *Direct Effect* | 21.17 | \$1,174,865 | \$2,124,467 | \$40,277 | \$246,234 |
| *Indirect Impacts* | 7.56 | \$330,930 | \$899,455 | \$37,926 | \$71,683 |
| *Induced Impacts* | 6.60 | \$270,289 | \$824,519 | \$52,928 | \$61,940 |
| ***Total Impact*** | **35.33** | **\$1,776,084** | **\$3,848,441** | **\$131,131** | **\$379,857** |
| ***Impact Multiplier*** | **1.67** | **1.51** | **1.81** | **3.26** | **1.54** |

**Table 3.**
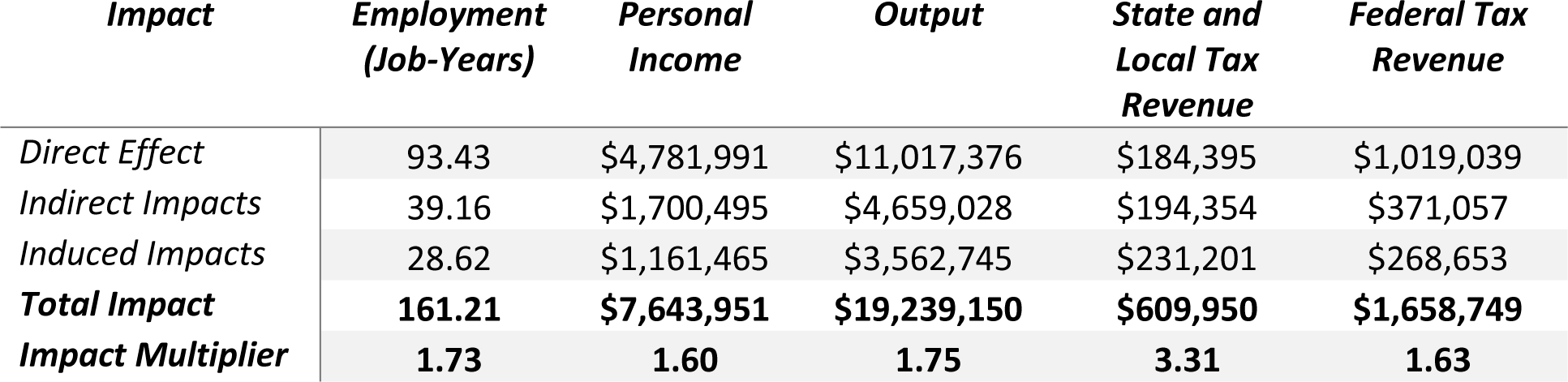
Cumulative Economic Impact of FCHES Federal Funding, 2016–2020

## RESULTS

### Economic Impacts

Table 1 shows annual NIH investment for FCHES. The combined annual NIH investment in FCHES reached $11 million.

Table 2 provides context to the overall assessment of the economic impact of the FCHES investment by examining the most recent completed year (2019) funded by NIH, which amounted to over $2.1 million.

This one year of investment was directly responsible for 21 jobs (expressed as years of full-time employment – job-years), over $1.1 million in personal income and over $286,000 in state/local and federal tax revenue (Table 2). Through the spending of the center, additional indirect impacts were generated in Genesee County, including more than 7 job-years, close to $331,000 personal income to employed persons, and almost $900,000 in additional economic output. In addition, the personal spending of the FCHES researchers and staff and of the employees in the indirectly created jobs generated induced impacts of close to 7 additional job-years, over $270,000 in personal income, and an economic output of over $800,000. Overall, combined direct, indirect, and induced impacts of one year of FCHES federal investment in Genesee County included over 35 job-years, close to $1.8 million in personal income, and more than $3.8 million in economic output. In addition, this combined economic activity generated over $500,000 in state/local and federal tax revenue.

Table 3 provides estimates of the economic impact of the total FCHES investment, which amount to approximately $11 million between 2016-2020. The overall federal investment in FCHES was directly responsible for 93 job-years, $4.8 million in personal income and over $1.2 million in state/local and federal tax revenue (Table 3). Additional indirect impacts were generated in Genesee County through the spending of the center, including 39 job-years, over $1.7 million personal income to employed persons, and close to $4.7 million in added economic output. In addition, the personal spending of the FCHES researchers and staff and of the employees in the indirectly created jobs generated induced impacts of close to 29 job-years, close to $1.2 million in personal income, and an economic output of over $3.5 million. Overall, combined direct, indirect, and induced impacts of the total FCHES federal investment in Genesee County included over 161 job-years, over $7.6 million in personal income, and more than $19.2 million in economic output. In addition, this combined economic activity generated close to $2.3 million in state/local and federal tax revenue. The impact multipliers show the ripple effect of the FCHES investment. For example, the overall output of over $19.2 million led to an impact multiplier of 1.75 – every $1 of federal FCHES investment led to an additional $.75 of economic output in Genesee County.

### Functional impacts

While the economic impact of FCHES was the focus of this study, the primary goal of FCHES is to address health disparities and chronic disease that cross boundaries and directly affect the Flint and Genesee County Community. These advancements in knowledge and practice are expected to benefit healthcare and health of the Flint and Genesee County population, and beyond. We refer to these benefits in the current study as *functional impacts*. The functional FCHES impacts include expanding the scientific knowledge by embracing broad and inclusive research themes, intervening to reduce physical and behavioral health disparities, improving methods in disparities research, improving practices through dissemination, developing networks of collaborators and stakeholders, increasing partner organization capacity to effectively participate in the research enterprise, and engaging and enhancing community residents awareness and participation in reducing health inequities.

As best examples of functional impacts, the FCHES research projects include the Church Challenge (CC) and Strengthening Flint Families (SFF). The Church Challenge objective is to support healthy living environments and improve the health of the Flint community, with a focus on effective blood pressure management. The CC was developed using community-based participatory approaches and is based on a church-based program developed by and for primarily African-American Flint church congregations. This multilevel intervention addresses health at the community (level 3), church (level 2), and individual (level 1) to reduce blood pressure, reduce chronic disease risk, and promote health equity and wellbeing in Flint. The CC RCT works within churches and with individuals and is designed to examine the effectiveness of a community-designed, community-based, multilevel physical activity and nutrition program relative to an enhanced treatment as usual - the Health and Wellness Program. By June 2020, the Church Challenge had increased food availability for 176 participants, connected 273 participants to local resources that support healthy living, and disseminated health improvement strategies through 21 churches.

Strengthening Flint Families is a community-based behavioral health intervention designed to reduce behavioral health disparities and improve family resilience in the Flint Community by providing multiple levels of peer support and family services. Strengthening Flint Families is comprised of three behavioral health interventions: at the Individual level: Peer Recovery Coaches (PRCs); at the Family level: Strengthening Families Program (SFP); and at the Community-level: a Multi-Media Campaign. Phase 1 of SFF consisted of conducting 87 organization assessments and 9 key information interviews to identify available resources, gaps in services, and barriers to accessing behavioral health services in Flint. Phase 2 of SFF is the implementation phase in which results from phase 1 are used to identify and implement referral sites, host sites, and adopter sites for the SFP and PRC services. SFF provided SFP referral information to 77 organizations and has established a dedicated phone line and email address for organizations and community members to make referrals to the SFP. At the individual level, 4 PRCs were certified and 2 focus groups were conducted with 10 local PRCs to better understand the community needs. At the family level, as of June 2020, 22 families were referred to the SFP and 4 cohorts of the SFP have been completed with 16 youth and 10 adults receiving the full SFP program. At the community level, SFF placed 3 bus bench ads in strategic locations in Flint in November 2019 to promote the SFP and general behavioral health and wellbeing. The Community Playlist radio show hosted by Kristen Senters Young (Community PI of SFF) has produced 98 episodes in the past year. The show promotes the SFP as well as provides additional positive messaging to the community on prevention, treatment, and recovery. Episodes average 22.8 plays on the SoundCloud platform with 193 plays as the highest number of plays for one episode. The SoundCloud platform has grown to 28 followers in the past few years. The SFF Facebook page has grown to 151 likes and 170 followers, the SFF Twitter page has grown to 61 followers, and the SFF Instagram page has grown to 91 followers in the past few years.

## DISCUSSION

Overall, combined direct, indirect, and induced impacts in Genesee County of the $11 million FCHES federal investment included over 161 job-years, over $7.6 million in personal income, and more than $19.2 million in economic output. In addition, this combined economic activity generated close to $2.3 million in state/local and federal tax revenue. The overall output of over $19.2 million led to an impact multiplier of 1.75 – every $1 of federal FCHES investment led to an additional $.75 of economic output in Genesee County.

This is the first study, to our knowledge, applying an I/O framework to estimate the economic impacts of a large multidisciplinary research center. The method may be relevant and generalizable to other TCCs or other large research centers such as FCHES. Our estimates of the FCHES economic impact are in line with the few prior studies assessing the economic impact of funded medical research. Our estimates, including the 1.75 output impact multiplier, are likely conservative as the economic impacts were estimated only for one Michigan county, assuming there were no spillover effects in the adjacent counties, or more broadly in Michigan. Chatterjee and DeVol (2012) evaluated economic returns of NIH funding in the Biosciences industry. They estimated that the funding led to an output impact multiplier of 1.70 of output in the bioscience industry. The long-term effects suggest impact multipliers as high as 3.20, depending on the model specification. United for Medical Research reported that in fiscal year 2018, total NIH research funds awarded in 50 states and D.C. was over $28 billion, with an estimated impact in total economic activity nationwide of close to $74 billion, which implies an impact multiplier of 2.63. Tripp and Grueber (2011) examined the economic impact of the Human Genome Project and used similar impact estimation method as this study. They found that the cumulative economic impact of Human Genome Sequencing federal funding (1998-2003) led to an output impact multiplier of close to 3. The study estimated the impact throughout the US economy, which may explain the larger multiplier compared to our estimated impact in one Michigan county.

In addition to the estimated economic impacts, the functional impacts included community-based multilevel interventions to improve health outcomes and equity in Flint, MI and Genesee County, MI. Other functional impacts included an enhanced understanding of transdisciplinary research, a community focus on gaps in health equity and commitment to equitable distribution of power and resources, enhanced capacity in conducting disparities research, improved practices through dissemination and implementation science, enhanced networks of collaborators and stakeholders, increased partner organization capacity to effectively participate in the research enterprise, and enhanced awareness and increased participation of community residents in reducing health inequities.

Limitations inherent to the use of IMPLAN and other I/O models include constant returns to scale, the assumption of no supply constraints, fixed input structure, and the static nature of the model. Strengths of our analyses include the fact that we used actual data on FCHES employment and compensation. As IMPLAN was designed to calculate the total I/O of an industry, it is unusual to have a complete set of employment and compensation data and therefore creates estimates of employment and compensation averages specific to the industry and the area under study. When we added actual FCHES employment and compensation to the model, we found that this not only significantly increased the precision of the model but increased the estimated overall impact of FCHES as well, mostly due to a higher number of job years created compared to the model’s estimated employment and compensation data. The larger impact in the specific model suggests that FCHES employs significantly more individuals per dollar of investment than the industry average in Genesee County.

## CONCLUSIONS

The FCHES research funding yields significant U.S. direct economic impacts above and beyond the direct NIH investment of $11 million. The total impact in Genesee County, MI, of the five-year NIH investment in FCHES was over $19 million, creating over 161 job-years, $7.6 million personal income, and close to $2.3 million in tax revenue. The economic impact estimation method may be relevant and generalizable to other TCCs or other large research centers such as FCHES. In addition, this study may be useful to government officials seeking to understand the direct and contemporary benefits of government investment in health research. Looking into the future, upcoming studies will assess more broadly the effects of the investment in FCHES in the counties bordering Genesee, MI, other counties, and statewide in Michigan.

## Data Availability

The data that support the findings of this study are available from IMPLAN, LLC but restrictions apply to the availability of these data, which were used under license for the current study, and so are not publicly available. Output data from the IMPLAN model are however available from the authors upon reasonable request and with permission of IMPLAN, LLC

## DECLARATIONS

### Ethics approval and consent to participate

Not Applicable

### Consent for publication

Not Applicable

### Competing interests

No competing interests.

### Funding

This project is supported by funding from the National Institute on Minority Health and Health Disparities (U54MD011227; PI Furr-Holden).

### Authors’ contributions

All authors approved the submitted version and agree both to be personally accountable for the author’s own contributions and to ensure that questions related to the accuracy or integrity of any part of the work, even ones in which the author was not personally involved, are appropriately investigated, resolved, and the resolution documented in the literature. CM and DFH led the conception of the study. CM led the design of the study, with substantial contributions from BWM. CM, BWM, CB, and LW drafted the study. CM, DFH, RE, EYL, STY, LV, and BJF substantively revised the study.

## Acknowledgements

The authors would like to thank IMPLAN, LLC for the use of their data and I/O model, and Brian Barlow for his guidance in the usage and interpretation of the data and models.

## Notes

### Competing Interest Statement

The authors have declared no competing interest.

### Author Declarations

The research reported in this study does not constitute human subject research

